# Genomic characterization of invasive typhoidal and non-typhoidal *Salmonella* in southwestern Nigeria

**DOI:** 10.1101/2022.01.28.22270011

**Authors:** Odion O. Ikhimiukor, Anderson O. Oaikhena, Ayorinde O. Afolayan, Abayomi Fadeyi, Aderemi Kehinde, Veronica O Ogunleye, Aaron O. Aboderin, Oyinlola O. Oduyebo, Charles J. Elikwu, Erkison Ewomazino Odih, Ifeoluwa Komolafe, Silvia Argimón, Abiodun Egwuenu, Ini Adebiyi, Oluwadamiloa A. Sadare, Tochi Okwor, Mihir Kekre, Anthony Underwood, Chikwe Ihekweazu, David M. Aanensen, Iruka N. Okeke

## Abstract

**Background:** Salmonellosis causes significant morbidity and mortality in Africa. Despite being endemic in Nigeria, information on circulating lineages of invasive *Salmonella* is sparse.

**Methods:** *Salmonella enterica* isolated from blood (n=60) and cerebrospinal fluid (CSF, n=3) between 2016 and 2020 from five tertiary hospitals in southwest Nigeria were antimicrobial susceptibility-tested and Illumina-sequenced. Genomes were analysed using publicly-available bioinformatic tools.

**Results:** Isolates and sequence types (STs) from blood were *S*. Typhi [ST1, n =1 and ST2, n =43] and invasive non-typhoidal *Salmonella* (iNTS) (*S. Enteritidis* [ST11, n=7], *S*. Durham [ST10, n=2], *S*. Rissen [ST8756, n=2], *S*. Chester [ST2063, n=1], *S*. Dublin [ST10, n=1], *S*. Infantis [ST603, n=1], *S*. Telelkebir [ST8757, n=1] and *S*. Typhimurium [ST313, n=1], *S*. Typhi ST2 (n=2) and *S*. Adabraka ST8757 (n=1) were recovered from CSF. Most *S*. Typhi belonged to genotype 3.1.1 (n=44), carried an IncY plasmid and had several antibiotic resistance genes (ARGs) including *bla*_TEM-1_ (n=38), *aph(6)-Id* (n=32), *tet(A)* (n=33), *sul2* (n=32), *dfrA14* (n=30) as well as quinolone resistance-conferring gyrA_S83Y SNPs (n=37). All *S*. Enteritidis harboured *aph(3’’)-Ib, bla*_TEM-1_, *catA1, dfrA7, sul1, sul2, tet(B)* genes, and a single ARG, *qnrB19*, was detected in *S*. Telelkebir.. Typhoidal toxins *cdtB, pltA* and *pltB* were detected in *S*. Typhi, Rissen, Chester, and Telelkebir.

**Conclusion:** Most invasive salmonelloses in south west Nigeria are vaccine-preventable infections due to multidrug-resistant, West African dominant Typhi lineage 3.1.1.. Invasive NTS serovars, including some harbouring typhoidal toxin or resistance genes represented a third of the isolates emphasizing the need for better diagnosis and surveillance.

**Author Summary:** Whole genome sequencing of 63 invasive *Salmonella* from 5 tertiary hospitals in Nigeria revealed multiple serovars including a dominant antibiotic-resistance-gene harbouring *S*. Typhi 3.1.1 genotype comprising a gyrA_S83Y and IncY plasmid. We also report invasive non-typhoidal *Salmonella* harbouring typhoidal toxins.

## INTRODUCTION

*Salmonella* are a group of Gram negative, motile, facultative anaerobic rod-shaped bacteria belonging to the *Enterobacteriaceae* family. This genus consists of two known species, *Salmonella enterica* and *Salmonella bongori. S. enterica* are further distributed across six subspecies, of which the *S. enterica* subsp. *enterica* are most reported in infections involving homeotherm animals [1]. Furthermore, *S. enterica* subsp. *enterica* consists of over 1500 serovars with distinct antigenic specificity [2]. The human host-adapted *S. enterica* subsp. *enterica* serovars are usually associated with three marked clinical syndromes: *Salmonella enterica* subsp. *enterica* serovar Typhi cause typhoid fever, and the non-typhoidal *Salmonella* (NTS) cause bacteraemia and gastroenteritis in immunocompromised (including persons with advanced HIV disease, cases of severe malaria and malnutrition in children) and immunocompetent persons, respectively [3,4]. *S*. Paratyphi A, B and C produce a syndrome similar to typhoid fever.

The public health impact of typhoidal and invasive non-typhoidal *Salmonella* infections is significant particularly in Africa and Asia where they have a great influence on morbidity and mortality [5,6]. For instance, an estimated 17.8 million cases of typhoid fever occur each year in low and middle-income countries (LMICs) [7]. An earlier estimate suggests that the burden of typhoid fever is >100 per 100000 individuals per annum in sub-Saharan Africa with an associated 1% mortality [8,9]. Furthermore, an estimated 26% (33,490 lives lost) of the annual global typhoid-related mortality is reported to occur in Africa [9]. The disease burden of typhoid in Nigeria is estimated at 364,791 typhoid cases resulting in 4,232 deaths (affecting 68% of individuals under 15 years of age) as at 2016 [10], however population-based data are only just becoming available [11]. Globally, NTS is estimated to cause approximately 94 million cases of gastroenteritis per annum worldwide with a resultant mortality of 155,000 [12]. In immunocompromised cases of the disease (amongst HIV-positive adults), NTS is reported to cause a 20% case fatality (212,000 deaths) in sub-Saharan Africa (SSA) annually, while also being responsible for over 1 million cases of bloodstream infections in children in SSA with a case fatality of 18.1% (197,000 child mortality) [4,13,14].

Although available reports suggest infection with *Salmonella enterica* to be the most common cause of bloodstream infections in Africa [15], the incidence and microbiology of typhoidal and invasive non-typhoidal *Salmonella* (iNTS) is still poorly understood. Many regions on the continent have garnered little or no attention in the literature [7]. Blood culture-based surveillance represents the standard method for assessing the epidemiology and aetiology of bacterial invasive infections [16]. Limited surveillance of invasive *Salmonella* on the Africa continent is majorly due to financial, logistical, and infrastructural constraints for the institution and maintenance of blood culture-based surveillance systems in the region [7,8,16,17].

Such limitations not only obscure the true burden and prevalence of invasive *Salmonella* infections in resource-limited settings but also limit opportunity for genomic surveillance of this pathogen. For instance, despite the huge burden of typhoid infections in Nigeria, before the current study, there were only 131 *Salmonella* genomes (all *S*. Typhi) from the country on Pathogenwatch (https://pathogen.watch/, 15th November 2021) [18], a web-based platform for surveillance of microbial genomes all of which were collected on or before 2013, and most from only two centres [19]. Lack of genomic surveillance information of invasive *Salmonella* in resource-limited countries, including Nigeria, may deter interventions necessary to ameliorate this burden, such as the typhoid conjugate vaccines [8,17,19,20]. Hence, this report provides genomic characterization of 2016-2020 invasive *Salmonella* retrieved from tertiary hospitals enrolled into Nigeria’s Antimicrobial Surveillance Network coordinated by the Nigeria Centre for Disease Control (NCDC).

## MATERIALS AND METHODS

### Ethical considerations

Isolates were obtained as part of the surveillance efforts in line with Nigeria’s national action plan. Ethical approval for using them in research was obtained from the University of Ibadan/University College Hospital ethics committee (UI/EC/15/093). Patient consent was not obtained and the data were analysed anonymously.

### Isolate collection, Identification and Antimicrobial Susceptibility Testing

Tertiary hospitals located in southwest Nigeria and enrolled into the Nigeria Antimicrobial Surveillance Network provided cryopreserved isolates from blood and cerebrospinal fluid to the AMR National reference laboratory. The isolates were from retrospectively batched periods of 2016-2018 (retrospective isolates), 2019 and 2020. The national reference lab in partnership with the Global Health Research Unit for the Genomic Surveillance of Antimicrobial Resistance (GHRU-GSAR) conducted the re-identification of the isolates using the Gram-negative (GN) test kit on (Ref: 21341) on VITEK 2 systems (version 2.0, Marcy-l’Etoile, France, Biomérieux). Briefly, the cryopreserved isolates (at −80 °C) are resuscitated before use for reidentification by subculturing onto Salmonella-Shigella Agar and incubated aerobically at 37°C. Isolated colonies from pure cultures are the streaked on Nutrient Agar (NA), incubated aerobically at 37°C. Isolated colonies on NA is then used to prepare inoculum for VITEK using GN cards. This test is based on forty-seven biochemical tests and a negative control. The cards contain wells with substrates for the different tests in dried form. The cards are inoculated with a saline suspension of the organisms before incubation. Upon incubation, biochemical reactions are read by the machine and recorded as positive or negative. A bionumber which is based upon the combination of different test results is then generated. The bionumber is compared to VITEK 2 robust database to match the organism and this is used to identify the organism. An added step for confirming identity of VITEK-identified isolates utilized whole-genome sequencing of all isolates. Antimicrobial susceptibility testing was done using VITEK AST N280 test kit (Ref: 413432). N280 Cards are incubated within the VITEK 2 compact upon inoculation with appropriate saline suspension of test organism. The minimum inhibitory concentration (MIC) is recorded as the highest concentration of an antibiotic for which no bacterial growth is observed. The MICs were interpreted as either resistant, intermediate or susceptible in accordance to CLSI standards [21].

### DNA extraction and Library preparation

The isolates were processed for the extraction of genomic DNA using Wizard DNA extraction kit (Promega; Wisconsin, USA) following manufacturer’s instructions. The extracted DNA was quantified on a Qubit fluorometer (Invitrogen; California, USA) using dsDNA Broad Range quantification assay. Double-stranded DNA libraries were prepared using the Covaris LC220 for fragmentation, and NEBNext Ultra II FS DNA library kit for Illumina with 384-unique indexes (New England Biolabs, Massachusetts, USA; Cat. No: E6617L). Libraries were sequenced on an Illumina HiSeq X10 (Illumina, California, USA).

### Genome Assembly

Generated sequence reads from Illumina runs were *de novo* assembled following GHRU protocols (https://gitlab.com/cgps/ghru/pipelines/dsl2/pipelines/assembly) using a Nextflow workflow which inclusively comprises of adapter trimming (trimmomatic v0.38), contamination detection (ConFindr v0.7.2), assembly (SPAdes v3.12.0), Quality Control (multiqc v1.7, qualifyr v1.4.4) and Bactinspector (v 0.1.3).

### Sequence typing of *Salmonella* genomes

Sequence reads were deposited in the *Salmonella* database on EnteroBase [22]. Multi-locus sequence types (MLST) for the isolates were determined and core-genome MLST calculated. Evolutionary relationship based on cgMLST of all *S*. Typhi of human origin from Africa deposited in Enterobase were determined [22]. The *Salmonella* genome assemblies were analysed using the *Salmonella* In-Silico Typing Resource (SISTR) for the prediction of serovars and serogroups (https://github.com/phac-nml/sistr_cmd). Genomes belonging to *S*. Typhi were loaded unto Pathogenwatch for the prediction of their genotypes [18].

### Identification of AMR, Plasmids, Virulence genes and *Salmonella* Pathogenicity islands

Determinants of AMR, virulence and plasmid replicons were identified following GHRU protocols (https://gitlab.com/cgps/ghru/pipelines). Prediction of *Salmonella* pathogenicity islands (SPIs) in the genomes was done by mapping raw reads to SPIs database (https://bitbucket.org/genomicepidemiology/spifinder_db)

### Single Nucleotide Polymorphism (SNP) calling and phylogeny

The sequence reads of the *S*. Typhi and *S*. Enteritidis genomes from our study were mapped to NCBI reference sequence, *Salmonella* enterica subsp. enterica serovar Typhi strain H12ESR00755-001A (assembly accession: GCF_001362195.2) and *Salmonella* enterica subsp. enterica serovar Enteritidis strain 18569 (assembly accession: GCF_000335875.2), respectively, to determine evolutionary relationship amongst the strains following GHRU nextflow SNP phylogeny protocols (https://gitlab.com/cgps/ghru/pipelines/snp_phylogeny).

Briefly, reads were trimmed (trimmomatic v0.38) and mapped to the reference genomes described above using bwa mem (v0.7.17) and variants were called and filtered using bcftools (v1.9). A pseudoalignment with the reference was used to generate a maximum likelihood tree using iqtree (v1.6.8) [23]. SNP distances between the genome pairs were calculated using snp-dists v.0.8.2 (https://github.com/tseemann/snp-dists) on the pseudo-genome alignment.

### Availability of sequence data

Raw sequence data generated from this study are deposited in the European Nucleotide Archive under Bioproject PRJEB29739. Accession numbers for each genome is available in Supplementary Table 1.

## RESULTS

### Invasive *Salmonella* from sentinel hospitals from Nigeria’s AMR Surveillance Network

Using the VITEK system for bacterial identification described above, a total of 69 isolates retrieved from patients from five (n=5) sentinel hospitals were identified as *Salmonella* spp., at the reference laboratory. However, results from whole-genome sequencing confirmed n=61 of these to be *Salmonella enterica*. In addition, two other isolates from our surveillance collection initially identified as *Escherichia coli* and *Acinetobacter baumanii* using VITEK were subsequently identified as *Salmonella enterica* using WGS.

The sixty-three (63) WGS-confirmed invasive *Salmonella* isolates have average number of contigs was 58 and N50 values ranged from 172132bp to 731013bp (average 246872 bp). The G+C (%) content of the genomes ranged from 51.86% - 52.37% (average 52.10%) (Supplementary Table 1). The isolates were retrieved from blood (n=60) and cerebrospinal fluid (n=3). The sending sentinel hospitals include: University of Ilorin Teaching Hospital (ILO, Ilorin, Kwara State, n=25), University College Hospital, Ibadan (UCH, Ibadan, Oyo State, n=23), Obafemi Awolowo University Teaching Hospital, Ile-Ife (OAU, Ile-Ife, Osun State, n=8), Lagos University Teaching Hospital (LUT, Idi-Araba, Lagos State, n=4) and Babcock University Teaching Hospital (BUT, Ilishan-Remo, Ogun State, n=3) (Fig. 1A). Majority of the isolates were retrieved in 2019 from ILO (n=22). Thirteen isolates had no year specified metadata but were retrospective isolates retrieved between 2016 and 2018 (Fig 1A). The hospitals are all in the southwestern part of Nigeria with ILO just north of the South-West geopolitical zone and all the rest within it. All the *Salmonella* isolates from cerebrospinal fluid were obtained from LUT.

**Fig 1:**
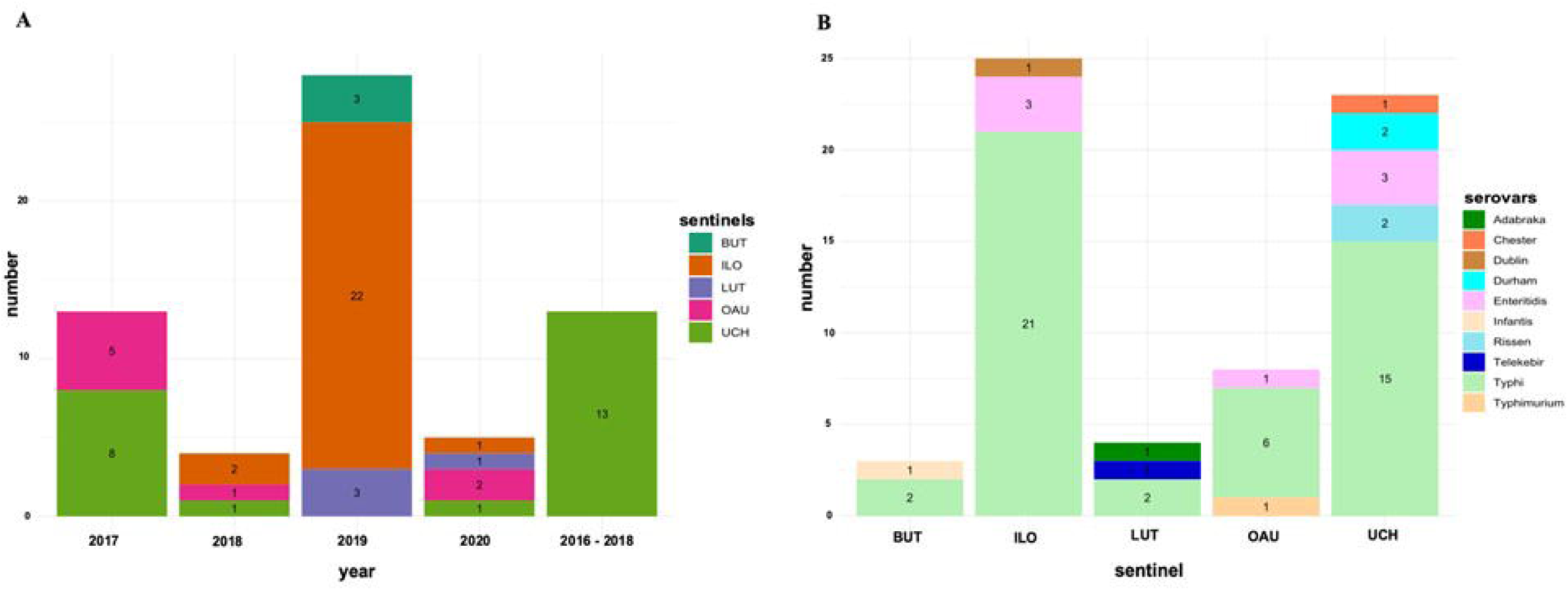
Epidemiological information showing: (A) number of Salmonella isolated received from the different sentinel hospitals at different years, and (B) Number of different *Salmonella* serotypes received from the different sentinel hospitals.

### Distribution of *Salmonella enterica* subsp. *enterica serovars* across sentinel hospitals

All the *Salmonella enterica* isolates belonged to the subspecies *enterica* but differed by serotype with a total of 10 serovars detected. They include Typhi (n=46), Enteritidis (n=7), Durham (n=2), Rissen (n=2), Adabraka (n=1), Chester (n=1), Dublin (n=1), Infantis (n=1), Telelkebir (n=1), Typhimurium (n=1). Three *Salmonella enterica* isolates belonging to serovars Adabraka (n=1) and Typhi (n=2) were retrieved from cerebrospinal fluid from LUT. All other *Salmonella* serovars were retrieved by blood culture at the respective sentinel sites (Supplementary Table 1). *Salmonella* Typhi and iNTS were recovered from all sentinel sites, with iNTS being much less frequently recovered (Fig. 1B).

### Sequence types, genotypes, and nucleotide polymorphisms

*Salmonella* sequence-typing based on Achtman’s MLST scheme [24] identified two *S*. Typhi Sequence Types (STs) (ST1 =1 and ST2 = 45). There were nine different iNTS STs. These included previously reported invasive STs: *S* Enteritidis ST11 (n=7) and *S* Typhimurium ST313 (n=1), which are repeatedly reported from Africa. Other iNTS were *S*. Dublin (ST10), *S*. Infantis (ST603), *S*. Durham (ST2010), *S*. Chester (ST2063), *S*. Telelkebir (ST2222). Two novel STs belonging to *S*. Rissen and *S*. Adabraka were curated and designated STs 8756 and 8757 respectively by EnteroBase.

To further place our *S*. Typhi genomes in a wider context, we performed cgMLST analysis based on differences in core genomes of our strains and all *S*. Typhi from human sources in Africa deposited in EnteroBase (n=980) (Fig 2). All genomes included in this study had similar core genome allelic differences at HC400, whereas at HC200 genomes from this study had similar allelic profile with 98.06% (n=961) of the genomes in the population. Genomes accounting for the difference in cluster numbers in the population at HC400 were from Nigeria (n=9, ∼0.92%), Cameroon (n=4, ∼0.41%) Algeria (n=3, ∼0.3%) Morocco (n=2, 0.2%) and Senegal (n=1, 0.1%). Generally, *S*. Typhi genomes from this study clustered with others from West Africa, including Nigeria, Cameroon, Togo, Mauritania, Mali, Burkina Faso, Guinea, Benin, and Ivory Coast, emphasizing further on their endemicity in the West Africa region (Fig 2).

**Fig 2:**
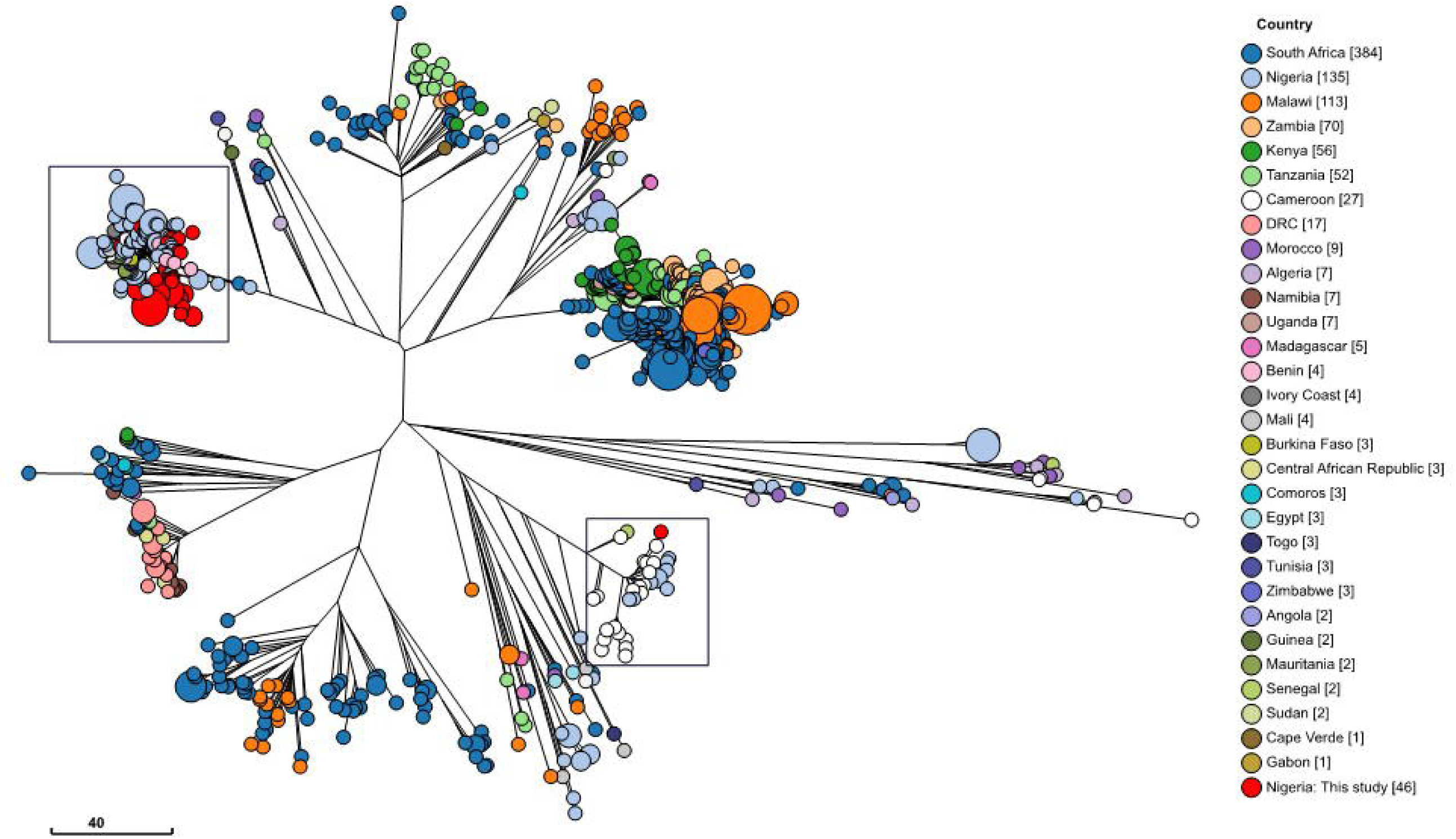
Grape tree showing core genome MLST of *S*. Typhi from human sources in Africa, deposited in the EnteroBase database. Red leaf labels are genomes from this study

Further, based on *S*. Typhi genotyping scheme, we observed that the isolate of *S*. Typhi ST1 belonged to genotype 4.1 (UCH), whereas genotypes 2.3.1 (n=1, UCH) and 3.1.1 (n= 44) were *S*. Typhi ST2 isolates. In addition, *S*. Typhi genomes from CSF (n=2) belonged to the 3.1.1 genotype. To further investigate the genetic relatedness of the genomes in this study, we report outcome of pairwise SNP differences among the genomes. Pairwise SNP difference ranged from 401 to 431 between *S*. Typhi 4.1 and other *S*. Typhi genotypes in this study, whereas it was 401 to 521 between *S*. Typhi 2.3.1 and other *S*. Typhi genotypes. The dominant Typhi genotype, *S*. Typhi 3.1.1 were 0 to 47 SNPs apart (Supplementary Table 3).. Among *S*. Enteritidis, pairwise SNP difference ranged from 0 to 44 in the three sentinel labs (UCH, OAU, ILO) where they were isolated. One pair of *S*. Enteritidis isolates from UCH were identical and the third isolate was 33 SNPs apart from them. The three *S*. Enteritidis isolates from ILO (n=3) were near identical having pairwise SNP range from 0 to 1 (Supplementary Table 4).

### Antimicrobial susceptibility profiles, antimicrobial resistance determinants and plasmids replicons in TS and NTS

Antimicrobial susceptibility testing revealed majority of the *S*. Typhi to be resistant to sulphamethoxazole/trimethoprim (SXT) and ampicillin (n=41 each) and nalidixic acid (n=36), of which three were ciprofloxacin non-susceptible, according to CLSI (2021) criteria. (Supplementary Table 2). While not relevant to the antimicrobial chemotherapy of invasive infections, resistance to nitrofurantoin was identified in the n=2 *S*. Typhi isolated from CSF and in n=9 isolates from blood with the highest MICs (128 μg/mL) seen in the CSF isolates only. Resistance to cephalosporins, cefuroxime and cefuroxime axetil was observed in *S*. Typhi 3.1.1 from UCH.

The single *S*. Typhi 2.3.1 isolate was resistant to ampicillin and SXT whereas no phenotypic resistance was observed with *S*. Typhi 4.1. All *S*. Enteritidis and *S*. Typhimurium were resistant to ampicillin and sulphamethoxazole/trimethoprim. Asides Telelkebir harbouring resistance to nalidixic acid and ciprofloxacin, other NTS were either susceptible or intermediately resistant to other antimicrobials (Supplementary Table 2). For example, *S*. Adabraka, Dublin and Telelkebir were intermediately resistant to cefuroxime axetil. (Supplemental Table 2 and microreact link for antimicrobial susceptibility testing: https://microreact.org/project/ahQ3Yb64nsbnhHMzz3WQn9-genomic-epidemiology-of-invasive-salmonella-in-southwestern-nigeria-ast-data)

A combined total of 14 acquired antimicrobial resistance genes (ARGs) conferring resistance to drugs within seven antibiotic classes were detected amongst the genomes. Amongst the *S*. Typhi genomes, n=36/46 harboured at least one ARG conferring reduced susceptibility to 5 antibiotic classes, with n=41 harbouring a sulphonamide resistance gene [*sul1* =9/46, *sul2* = 33/46)] and n=39 each harbouring a beta-lactam (*bla*_TEM-1_), tetracyclines (*tetA*, n=33 and *tetB*, n=6), and trimethoprim resistance determinant (*dfrA1*, [n=1], *dfrA15* [n=8] and *dfrA14* [n=30]). In addition, chloramphenicol resistance genes, *catA1*, were also detected in the genomes (n=8). Point mutations identified among the sequenced *S*. Typhi isolates were those associated with the quinolone resistance determining region (QRDR), gyrA_S83Y SNPs (n=37), which mediate resistance to fluoroquinolones (Fig 3). Furthermore, n=45 of the *S*. Typhi genomes had at least one plasmid predicted to occur in each genome. Majority (n=33) possessed an IncY plasmid replicon, plasmid replicons IncFIA_HI1, IncHIA and IncHIB were respectively detected in n=9 of *S*. Typhi genomes whereas one isolate harboured an IncQ plasmid replicon (Fig 3).

**Fig 3:**
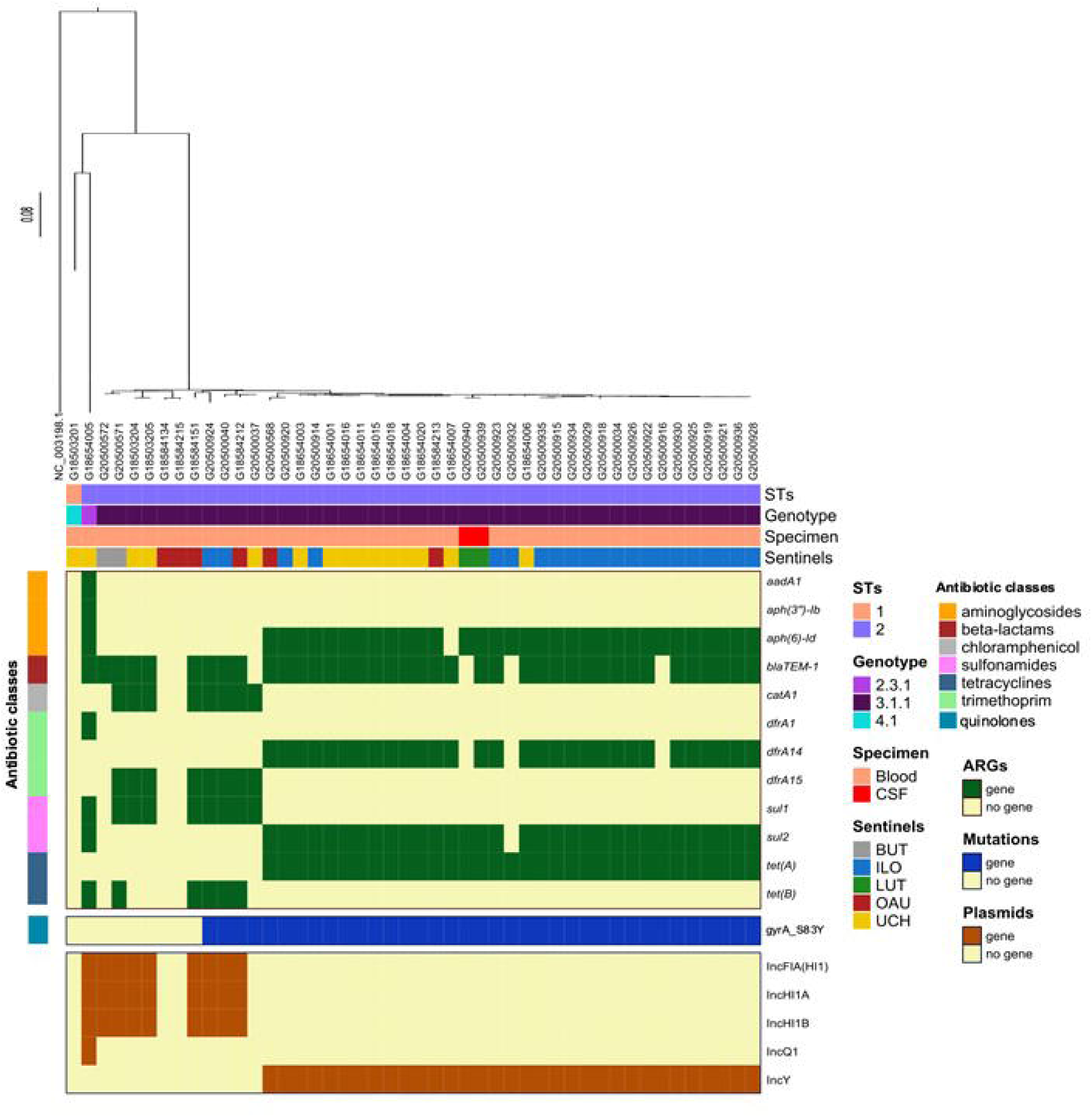
SNP-phylogeny based tree and gene presence/absence showing the genomic profile of *Salmonella* Typhi genomes retrieved from 5 sentinel laboratories in Nigeria. ARGs: antibiotic resistance genes, STs: Sequence types

For the iNTS, *S*. Enteritidis genomes possessed at least one ARG to six antibiotic classes. All Isolates of this serotype harboured *aph(3”)-Ib, bla*_TEM-1_, *catA1, dfrA7, sul1, sul2, tet(B)* genes, and only differed in the absence/presence of *aph(6)-Id* (n=4) (Fig 4). In tandem, *S*. Typhimurium harbour ARGs [*aadA1, aph(3’’)-Ib, aph(6)-Id, bla*_TEM-1_, *catA1, dfrA1, sul1, sul2*] encoding resistance to 5 antibiotic classes (Fig 5). The only occurring quinolone resistance gene among isolates in this study, *qnrB19*, was detected in *S*. Telelkebir. No ARGs were detected in *Salmonella* serovars Chester, Rissen, Durham, Infantis, Adabraka and Dublin. Antimicrobial point mutations identified among iNTS were associated with *gyrA* and *parC* gene regions (Fig 5). The quinolone resistance conferring gyrA_D87Y SNPs were identified only amongst *S*. Enteritidis (ILO, n = 3 and UCH, n = 1), whereas the parC_T57S mutations were detected in all iNTS except *S*. Enteritidis and *S*. Typhimurium. Plasmids were predicted to occur only in *S*. Dublin [IncFII(S), IncX1 and IncX1_1], *S*. Enteritidis (IncI1 and IncQ1), *S*. Typhimurium [IncFIB, IncFII(S) and IncQ1] among the iNTS.

**Fig 4:**
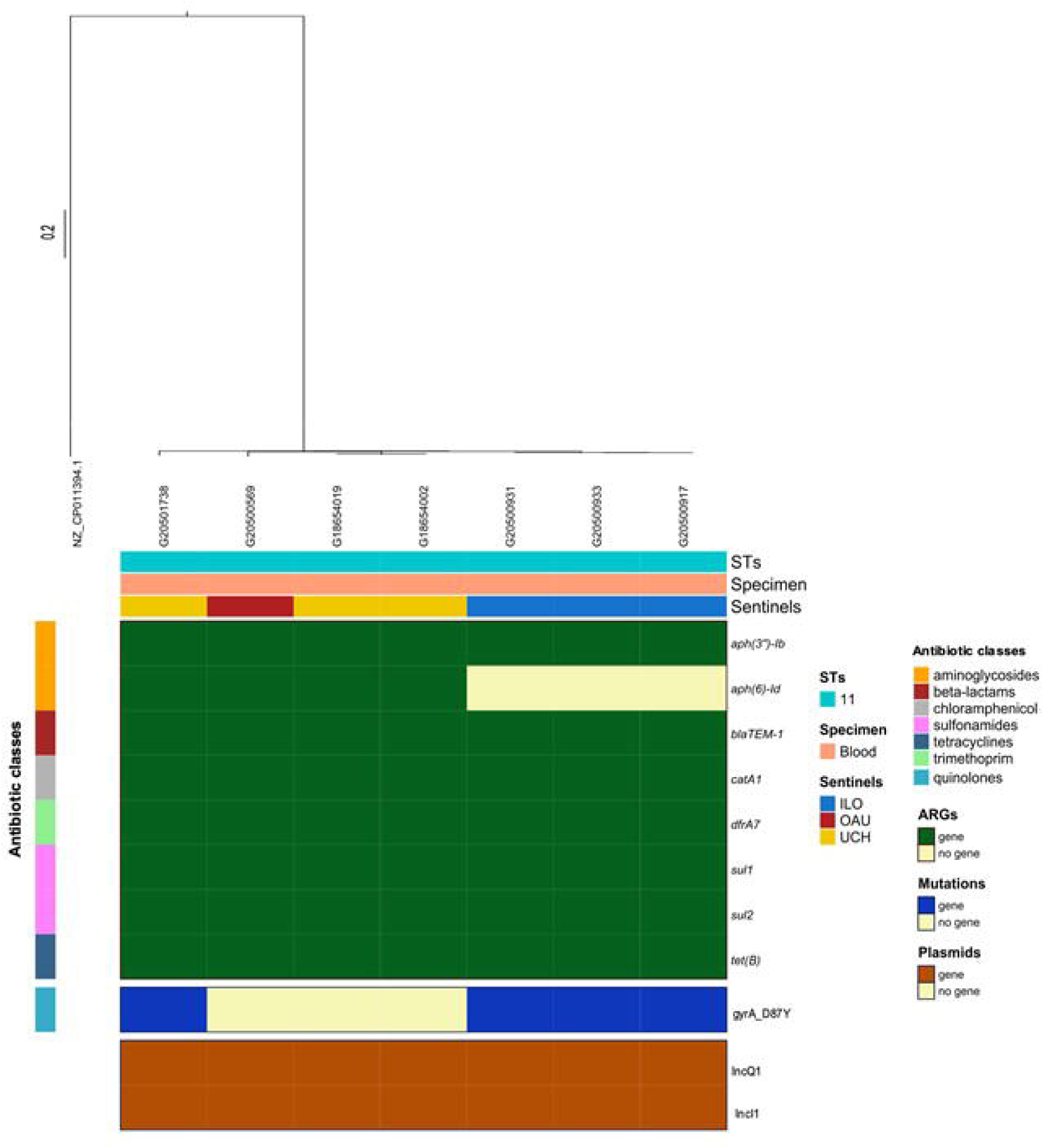
SNP-phylogeny based tree and gene presence/absence map showing the genomic profile of *Salmonella* Enteritidis retrieved from 3 sentinel laboratories in Nigeria. ARGs: antibiotic resistance genes, STs: Sequence types.

**Fig 5.**
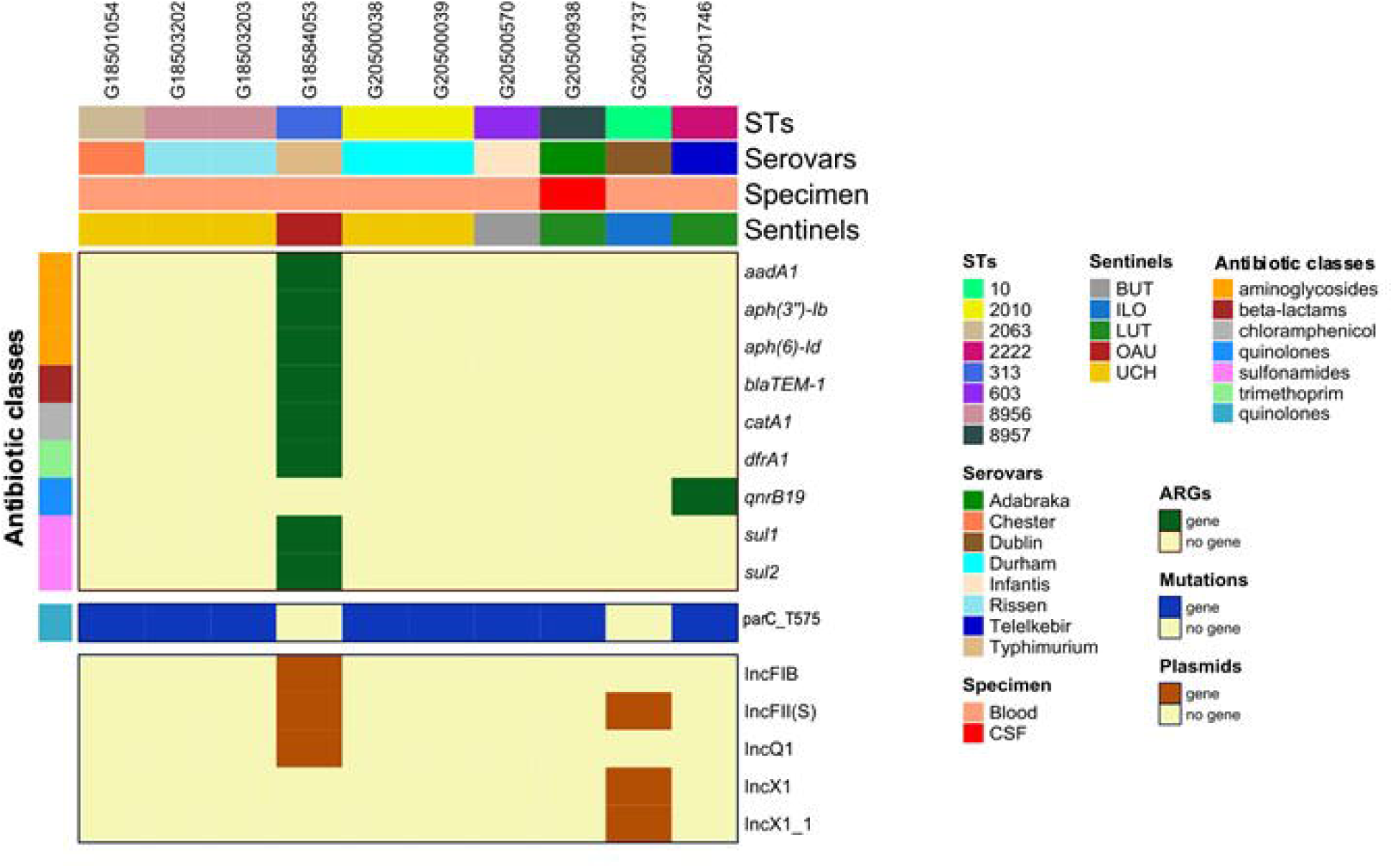
Gene presence/absence map showing the genomic profile of non-typhoidal *Salmonella* retrieved from 5 sentinel laboratories in Nigeria. ARGs: antibiotic resistance genes, STs: Sequence types

### Predominant IncY + gyrA_S83Y + *tetA* harbouring *S*. Typhi 3.1.1 in Nigeria

We observe that all *S*. Typhi of the 3.1.1 lineage harbouring an IncY plasmid replicon (n=33/46) similarly possessed the gyrA_S83Y chromosomal gene mutation and harboured a *tetA* gene. Although other antimicrobial resistance genes were seen at slightly lower numbers (n= 32/33 for *sul2* and *aph(6)-*Id and n=30/33 for *bla*_TEM-1_ and *dfrA14*), the IncY+gyrA_S83Y+*tetA* in *S*. Typhi 3.1.1 phenomenon was observed to occur in all the sentinel hospitals in this study. Additionally, maximum pairwise SNP distance between the variants in this cluster was 23. This is approximately twice as less of what was determined in *S*. Typhi 3.1.1 outside this cluster (n=47), thereby, emphasizing clonality within this cluster.

### *Salmonella* virulence determinants and predicted pathogenicity islands

The isolates possessed a plethora of virulence determinants (Supplementary Table 2). Among the *S*. Typhi genomes, a total of 98 virulence determinants were detected, and 97 of these were conserved within members of this serovar (with the exception of *pipB2* gene in an *S*. Typhi 3.1.1 from UCH).

A total of 106 virulence genes were detected among the *S*. Enteritidis genomes, 104 of these were conserved within these genomes, with 2 strains from UCH lacking either a *Salmonella* secreted protein H (*sspH*) or secretion system effector I (*sseI*). A total of 122 virulence genes were detected in the iNTS genomes, and n=86 of these were conserved in all iNTS genomes. For instance, the iNTS possessed genes encoding (i) Adherence; such as *agf* – thin aggregative fimbrae or curli (*csgABCDEGF*), *misl* – an autotransporter protein, *pef* - plasmid-encoded fimbrae (present only in *S*. Typhimurium) *ratB* (carried by iNTS strains harbouring CS54 islands), *shdA* (only found in *S*. Infantis), *sinH* (detected in all NTS except *S*. Enteritidis) and Type 1 fimbrae (*fimCDFHI*) (ii) Stress adaptation; *sodCI* – superoxide dismutase (detected in iNTS serovars except Durham, Chester and Rissen and Infantis and Telelkebir), *sopA* (not detected in *S*. Infantis) (iii) Nutritional/metabolic factor (*mgtBC*, present in all strains) (iii) Antimicrobial activity/competitive advantage; such as macrophage inducible genes (*mig-14*, present in all strains) and (iv) Enterotoxin; T3SS effectors – *spvBC* (in *S*. Typhimurium, Enteritidis and Dublin), *avrA* (in all iNTS except *S*. Dublin) and Typhoidal toxin - *cdtB* (present in *S*. Durham, *S*. Telelkebir and *S*. Chester).

Since the *cdtB* are reported to be co-located with other cytolethal distending toxins (*cdt*), pertussis-like toxins A (*pltA*) and B (*pltB*), on same pathogenicity islet [25], we ran a blast search of our strains for the presence of *pltA* and *pltB*. The nucleotide sequences were extracted from the virulence factor database (VFDB) and used as a local database for a blast search against our iNTS genomes. Our results reveal high similarity (100% coverage and ≥ 96.62% identity) with *cdtB, pltA* and *pltB* genes in the iNTS genomes (*S*. Chester, *S*. Durham and *S*. Telelkebir).

Eleven and twelve *Salmonella* pathogenicity islands (SPIs) were predicted in *S*. Typhi and iNTS genomes, respectively (Fig 6). All *S*. Typhi were predicted to have 10 SPIs, i.e., SPI-1, SPI-2, SPI-3, SPI-4, SPI-5, SPI-6, SPI-7, SPI-8, SPI-9, SPI-10 and SPI-12. However, SPI-4 was predicted to occur only in *S*. Typhi lineages 2.3.1 and 4.1. In contrast to *S*. Typhi, only SPI-3 was predicted to occur in all the iNTS genomes. Certain SPIs were shown to be associated with members of certain serovars. For instance, SPI-2 and SPI-8 were detected only in *S*. Typhimurium and *S*. Rissen, respectively. Other pathogenicity islands were detected in this study (Fig 3), such as SPI-4 (*S*. Adabraka, Chester, Typhimurium), SPI-6 (all NTS except *S*. Durham, Rissen and Telelkebir), SPI-12 (all NTS except S. Chester, Durham, Rissen and Telelkebir) and CS54_island was detected in *S*. Dublin, *S*. Typhimurium, *S*. Infantis and *S*. Enteritidis (n=6).

**Fig 6:**
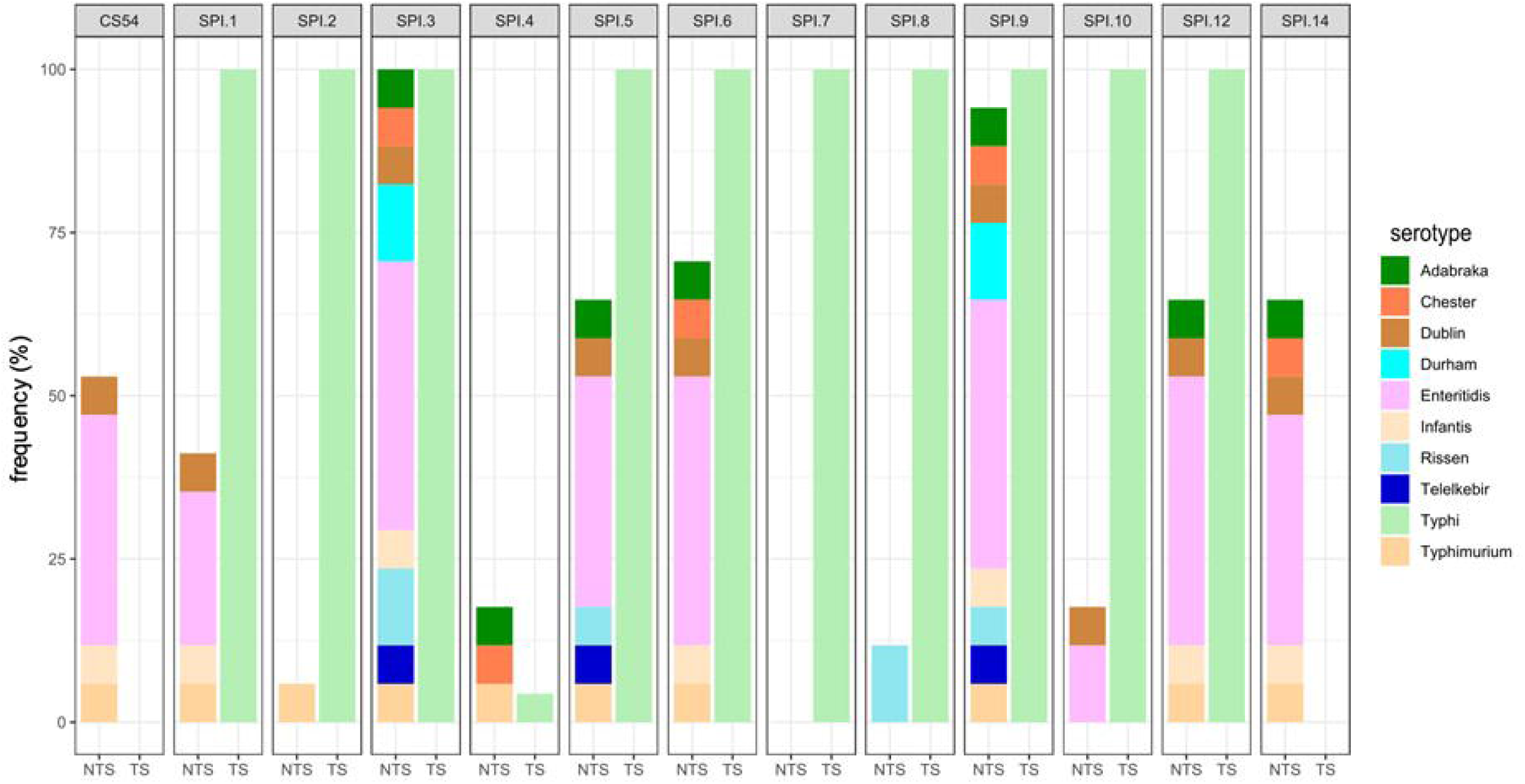
Frequency of occurrence of *Salmonella* pathogenicity island in TS (Typhoidal Salmonella) and NTS (Non-typhoidal Salmonella) in this study.

## DISCUSSION

In this report we present the outcome of genomic characterization of invasive *Salmonella* infections from AMR surveillance in sentinel hospitals in Nigeria. The genomic characterization of invasive *Salmonella* isolates in this study was possible because these hospitals perform blood culture and are enrolled in Nigeria’s new antimicrobial resistance surveillance system, which offers genomic services at the National Reference Laboratory level [26]. Nonetheless, blood culture is available at very few institutions in Nigeria, a limitation still prevalent in many African settings that impacts the genomic surveillance of invasive *Salmonella*. [8,16,27]. Moreover, these sentinels perform very few blood cultures so that the isolates studied here represent a very small proportion of circulating strains.

Using high-throughput WGS and bioinformatic analytics, we were able to determine prevalent serotypes and dominant genotypes of invasive *Salmonella* infections. Most of the *Salmonella* isolates cultured from blood were *S*. Typhi, as has been previously reported from different parts of Nigeria, including Ibadan [28–30], even though iNTS may predominate in some other African settings [3,31]. While our data are few, the predominance of Typhi at all sites points to a significant burden of severe disease that could be averted if Typhoid Conjugate Vaccines were deployed in Nigeria. Out of a total of 10 *S*. Typhi genotypes recorded from Nigeria in Pathogenwatch, three were identified in this study. The *S*. Typhi genotype 3.1.1 we report was similarly common in the Nigeria cluster on Pathogenwatch (n=87/131). As in our study, this cluster possessed similar prevalence of genetic determinants of beta-lactam (*bla*_TEM-1_ – 83.90%) resistance, indicating that these determinants are well-conserved in the genotype. However, prevalence of other AMR genetic determinants from this lineage such as *catA1, sul1, sul2, dfrA14, dfrA15, tetA, tetB* were similar with what is reported from this study, but at different rates.

This multidrug-resistance gene-encoding *S*. Typhi 3.1.1 is shown to be one of the broadest lineages in sub-Saharan Africa and endemic in the West Africa region [19,32,33]. This *S*. Typhi genotype is frequently reported to multidrug and ciprofloxacin resistant [29,31]. All S. Typhi 3.1.1 (except one from OAU) harbouring quinolone-conferring SNPs in *gyrA* showed phenotypic resistance to nalidixic acid. Additionally, we observed that the *S*. Typhi genotype 3.1.1 clone variants harboured an IncY + gyrA_S83Y + *tetA* genes. The Pathogenwatch database includes three *S*. Typhi 3.1.1 strains isolated from blood samples in 2013, in Abuja, north-central capital of Nigeria with similar clonal characteristics (having same genotype, plasmid replicon, chromosomal QRDR and antimicrobial resistance gene, *tetA*) [19]. Outside Nigeria, this lineage has also been identified in the United Kingdom (accession: SRR7165434, SRR5585020) [32]. Our data suggest that this resistant sub-lineage is predominant in our setting and should be sought elsewhere in Nigeria and the region. In addition, long read sequencing to unveil the carriage of the IncY plasmids would be potentially vital to understanding the success of this lineage in Nigeria.

*S*. Enteritidis were the most frequently recovered iNTS in our study and sent from three sentinel hospitals. This outcome contrasts with earlier reports of *S*. Typhimurium ST313 as a predominant serotype in eastern and southern Africa, but also present across the continent, including Nigeria [15,34,35], but it is concordant with more recent reports describing *S*. Enteritidis in higher proportions in invasive infections in The Gambia [36]. Several of the *S*. Enteritidis in our study were multidrug resistant (resistant to ampicillin, SXT, nalidixic acid). This multidrug resistant clone has also been reported in bacteraemia in other parts of Africa [37,38]. We observed that *S*. Enteritidis retrieved from different patients in ILO (in 2019) had highly similar genetic features (antimicrobial resistance determinants, virulence, plasmids replicons) and clustered together at 0-1 SNP distances between them. The isolates were recovered on the 27^th^ of June, 26^th^ of August and 28^th^ of August 2019 and their genetic, geographic and temporal connectedness may be indicative of a previously unrecognized outbreak. Both *S*. Typhimurium ST313 and *S*. Enteritidis ST11 are dominant, human-adapted clones in sub-Saharan Africa [38] and are a major cause of invasive disease, with a corresponding high case-fatality rate [14]. These serovars are justifiably vaccine development priorities. Non typhoidal *Salmonella* serovars, such as Dublin, Infantis, Chester, Rissen have been reported severally from food animals [39–43], their presence in human invasive human infection may attest to concerns with water and food safety, including animal contact [44–46]. Although no ARGs were detected in this genome of these serovars, they remain a public health concern [46,47]. The single occurrence of an acquired quinolone resistance gene, *qnrB19* in this study was detected in *S*. Telelkebir. The strain also harboured phenotypic resistance to the quinolone, nalidixic acid and ciprofloxacin. *S*. Telelkebir has been reported a few times in Africa (as seen in Enterobase, [48]), and are more commonly reported in parts of Europe, China and USA [49]. The expansion of atypical *Salmonella* serovars in invasive infections is associated with a high health burden [17,36]. Invasive NTS vaccines in the pipeline may not cover all NTS serovars [50], and we identified several in this study, harbouring an assortment of virulence and antimicrobial resistance determinants. This points to the need for widespread and robust access to invasive *Salmonella* diagnostics in Nigeria to elucidate on the burden and make a case for serovar vaccine priorities [17,36,49].[49][17,36]

Amongst a plethora of virulence determinants present on both *S*. Typhi and iNTS, we observed that *S*. Telelkebir, *S*. Durham and *S*. Chester isolates harboured the cytolethal distending toxin islet genes (*cdtB, pltA, pltB*) also known as typhoid toxin. These toxins were originally thought to be restricted to serovars Typhi and Paratyphi A [51]. However, these have now been reported in other NTS serovars including Bredeney, Javiana, Montevideo, Schwarzengrund, and more recently in Telelkebir [52–54]. A literature search on PubMed and Google Scholar revealed little information on these toxins being reported in *S*. Durham and *S*. Chester. The cytolethal distending toxin islet cause DNA damage and cell cycle arrest in impaired cells [55]. More implicatively, these genes encoded by NTS serovars have been reported to play vital roles in disease pathogenesis [53,54]. Many of *Salmonella* virulence determinants are clustered in pathogenicity island on the bacterial chromosome, playing key roles in disease pathogenesis [56,57]. A variety of SPIs were identified in this study. The SPI-7 which were exclusively detected in *S*. Typhi in this study are known to be large and major backbone constituent of *S*. Typhi, harbouring several virulence determinants including the Vi antigen [58]. Like in this study, the CS54 island and SPI-14 island are more commonly detected NTS [59–62], with scarce reports in *S*. Typhi, and the CS54 island are suggested to have evolved over multiple horizontal transfers [63]. Thus, this study emphasizes on an expanding number of serovars causing invasive infections in the country, and the public health implications therein. Further studies focussed on molecular analysis of gene content of SPIs in invasive *Salmonella* infections could be pertinent in understanding pathogenesis and aid in the advancement of treatment options [64].

## CONCLUSION

The outcome of our study emphasizes the need for expanded genomic surveillance of invasive *Salmonella* infections in Nigeria as a valuable tool to monitor antibiotic resistance spread and genetic characterization of circulating lineages in Nigeria. Close monitoring of the dominant *S*. Typhi 3.1.1 clone harbouring the IncY plasmid replicon and gyrA_S83Y chromosomal mutation, identified in all the tertiary hospitals in this study, including other serovars is vital, and this may help to establish strategies for empirical treatment and control of spread of antibiotic resistant lineages. Furthermore, our data suggests that introducing typhoid conjugate vaccines, recommended by the World Health Organization for countries like Nigeria that have a high typhoid disease burden, will have a significant impact on health [65]. Development of vaccines which target NTS would be useful in reducing the overall burden of NTS on the continent. Rigorous surveillance plays an essential part in determining which serovars most require coverage, as we observe *S*. Enteritidis to be most prevalent NTS in invasive infections in southwest Nigeria, and hence recommended as vaccine priorities. Importantly, broader protective effects may be achieved by improvements in water, sanitation and hygiene that could interrupt transmission of the causes of typhoid and other invasive salmonellosis.

## Data Availability

All data produced in the present work are contained in the manuscript and its posted supplementary data.

## FUNDING

This work was supported by Official Development Assistance (ODA) funding from the National Institute of Health Research https://www.nihr.ac.uk/ (grant number 16_136_111) to DMA and INO and the Wellcome Trust https://wellcome.org/ grant number 206194 to DMA. INO was an African Research Leader supported by the UK Medical Research Council (MRC) https://mrc.ukri.org/ and the UK Department for International Development (DFID) under the MRC/DFID Concordat agreement that is also part of the EDCTP2 program supported by the European Union (Award # MR/L00464X/1) and is presently a Calestous Juma Science Leadership Fellow supported by the Bill and Melinda Gates Foundation https://www.gatesfoundation.org/ (Award # INV-036234). The funders had no role in study design, data collection and analysis, decision to publish, or preparation of the manuscript.

## ACKNOWLEDGEMENT

We thank Chinenye Ekemezie, Ifeoluwa J. Akintayo, Dorothy U Cyril-Okoh, Abeeb A. Adeniyi and Ifeanyi E. Mba for excellent technical contributions and Jola-Ade J. Ajiboye for logistic assistance. We are grateful to the administration and staff of the five sentinels for supporting them in surveillance.

## Notes

### Competing Interest Statement

The authors have declared no competing interest.

### Author Declarations

Ethical approval for research using these isolates was obtained from the University of Ibadan/University College Hospital institutional review board

### Summary of Updates

Antimicrobial susceptibility testing data has been included and the figures have been refined. Edits were also made to the text.

## REFERENCES

1. Giner-Lamia J, Vinuesa P, Betancor L, Silva C, Bisio J, Soleto L, Chabalgoity JA, Puente JL, Soncini FC, García-Vescovi E, Flores G, Pedraza J, Yim L, García C, Astocondor L, Ochoa T, Hinostroza N, Graciela Pucciarelli M, Hernández-Alvarez A, Del Moral V, García-Del Portillo F. Genome analysis of Salmonella enterica subsp. diarizonae isolates from invasive human infections reveals enrichment of virulence-related functions in lineage ST1256. BMC Genomics. 2019 Jan 31;20(1):1–14.

2. Issenhuth-Jeanjean S, Roggentin P, Mikoleit M, Guibourdenche M, De Pinna E, Nair S, Fields PI, Weill FX. Supplement 2008–2010 (no. 48) to the White–Kauffmann–Le Minor scheme. Research in Microbiology. 2014 Sep 1;165(7):526–30.

3. Feasey NA, Dougan G, Kingsley RA, Heyderman RS, Gordon MA. Invasive non-typhoidal salmonella disease: an emerging and neglected tropical disease in Africa. The Lancet. 2012 Jun 30;379(9835):2489–99.

4. Keestra-Gounder AM, Tsolis RM, Bäumler AJ. Now you see me, now you don’t: the interaction of Salmonella with innate immune receptors. Nature Reviews Microbiology 2015 13:4. 2015 Mar 9;13(4):206–16.

5. Smith SI, Seriki A, Ajayi A. Typhoidal and non-typhoidal Salmonella infections in Africa. Vol. 35, European Journal of Clinical Microbiology and Infectious Diseases. 2016. p. 1913–22.

6. Phu Huong Lan N, Le Thi Phuong T, Nguyen Huu H, Thuy L, Mather AE, Park SE, Marks F, Thwaites GE, Van Vinh Chau N, Thompson CN, Baker S. Invasive Non-typhoidal Salmonella Infections in Asia: Clinical Observations, Disease Outcome and Dominant Serovars from an Infectious Disease Hospital in Vietnam. PLoS Neglected Tropical Diseases. 2016 Aug 11;10(8):e0004857.

7. Antillón M, Warren JL, Crawford FW, Weinberger DM, Kürüm E, Pak GD, Marks F, Pitzer VE. The burden of typhoid fever in low-and middle-income countries: A meta-regression approach. PLOS Neglected Tropical Diseases. 2017 Feb 27;11(2):e0005376.

8. Crump JA, Heyderman RS. A Perspective on Invasive Salmonella Disease in Africa. Clinical Infectious Diseases. 2015 Nov 1;61(Suppl_4):S235–40.

9. Mogasale V, Maskery B, Ochiai RL, Lee JS, Mogasale V V., Ramani E, Kim YE, Park JK, Wierzba TF. Burden of typhoid fever in low-income and middle-income countries: a systematic, literature-based update with risk-factor adjustment. The Lancet Global Health. 2014 Oct 1;2(10):e570–80.

10. TyVAC. Potential of typhoid conjugate vaccines in Nigeria. 2017. 2017 Nov.

11. Park SE, Toy T, Cruz Espinoza LM, Panzner U, Mogeni OD, Im J, Poudyal N, Pak GD, Seo H, Chon Y, Schütt-Gerowitt H, Mogasale V, Ramani E, Dey A, Park JY, Kim JH, Seo HJ, Jeon HJ, Haselbeck A, Conway Roy K, MacWright W, Adu-Sarkodie Y, Owusu-Dabo E, Osei I, Owusu M, Rakotozandrindrainy R, Soura AB, Kabore LP, Teferi M, Okeke IN, Kehinde A, Popoola O, Jacobs J, Lunguya Metila O, Meyer CG, Crump JA, Elias S, MacLennan CA, Parry CM, Baker S, Mintz ED, Breiman RF, Clemens JD, Marks F. The Severe Typhoid Fever in Africa Program: Study Design and Methodology to Assess Disease Severity, Host Immunity, and Carriage Associated with Invasive Salmonellosis. Clinical Infectious Diseases. 2019 Oct 30;69(Supplement_6):S422–34.

12. Majowicz SE, Musto J, Scallan E, Angulo FJ, Kirk M, O’Brien SJ, Jones TF, Fazil A, Hoekstra RM. The global burden of nontyphoidal salmonella gastroenteritis. Clinical Infectious Diseases. 2010 Mar 15;50(6):882–9.

13. Gordon MA. Salmonella infections in immunocompromised adults. Journal of Infection. 2008 Jun 1;56(6):413–22.

14. Reddy EA, Shaw A V., Crump JA. Community-acquired bloodstream infections in Africa: a systematic review and meta-analysis. The Lancet Infectious Diseases. 2010 Jun 1;10(6):417–32.

15. Marchello CS, Dale AP, Pisharody S, Rubach MP, Crump JA. A Systematic Review and Meta-analysis of the Prevalence of Community-Onset Bloodstream Infections among Hospitalized Patients in Africa and Asia. 2019;

16. Ombelet S, Barbé B, Affolabi D, Ronat JB, Lompo P, Lunguya O, Jacobs J, Hardy L. Best Practices of Blood Cultures in Low- and Middle-Income Countries. Frontiers in Medicine. 2019 Jun 18;6.

17. Balasubramanian R, Im J, Lee JS, Jeon HJ, Mogeni OD, Kim JH, Rakotozandrindrainy R, Baker S, Marks F. The global burden and epidemiology of invasive non-typhoidal Salmonella infections. https://doi.org/101080/2164551520181504717. 2018 mJun 3;15(6):1421–6.

18. Argimón S, Yeats CA, Goater RJ, Abudahab K, Taylor B, Underwood A, Sánchez-Busó L, Wong VK, Dyson ZA, Nair S, Park SE, Marks F, Page AJ, Keane JA, Baker S, Holt KE, Dougan G, Aanensen DM. A global resource for genomic predictions of antimicrobial resistance and surveillance of Salmonella Typhi at pathogenwatch. Nature Communications 2021 12:1 [Internet]. 2021 May 17 [cited 2021 Nov 13];12(1):1–12. Available from: https://www.nature.com/articles/s41467-021-23091-2

19. Wong VK, Holt KE, Okoro C, Baker S, Pickard D, Marks F, Page AJ, Olanipekun G, Munir H, Alter R, Fey PD, Feasey NA, Weill FX, le Hello S, Hart PJ, Kariuki S, Breiman RF, Gordon MA, Heyderman RS, Jacobs J, Lunguya O, Msefula C, MacLennan CA, Keddy KH, Smith AM, Onsare RS, de Pinna E, Nair S, Amos B, Dougan G, Obaro S, Parkhill J, Kingsley RA, Thomson NR, Keane JA, Hawkey J, Edwards DJ, Dyson ZA, Harris SR, Cain AK, Hadfield J, Klemm EJ, Watson CH, John Edmunds W, Chabalgoity JA, Kama M, Jenkins K, Dutta S, Campos J, Thompson C, Dolecek C, Parry CM, Karkey A, Kim Mulholland E, Campbell JI, Dongol S, Basnyat B, Arjyal A, Dufour M, Bandaranayake D, Toleafoa TN, Pravin Singh S, Hatta M, Isaia L, Thwaites G, Turner P, Soeng S, Crump JA, Nille EJ, Thanh DP, Valcanis M, Powling J, Dimovski K, Hogg G, Connor TR, Dave J, Murphy N, Holliman R, Sefton A, Millar M, Farrar J, Mather AE. Molecular Surveillance Identifies Multiple Transmissions of Typhoid in West Africa. PLoS Negl Trop Dis [Internet]. 2016 Sep 22 [cited 2022 May 2];10(9). Available from: https://pubmed.ncbi.nlm.nih.gov/27657909/

20. Carey ME, Steele AD. The Severe Typhoid Fever in Africa Program Highlights the Need for Broad Deployment of Typhoid Conjugate Vaccines. Clinical Infectious Diseases. 2019 Oct 30;69(Supplement_6):S413–6.

21. Clinical and Laboratory Standards Institute (CLSI). Performance Standards for Antimicrobial Susceptibility Testing. 2021, 31st ed, CLSI supplement M100.

22. Zhou Z, Alikhan NF, Mohamed K, Fan Y, Group the AS, Achtman M, Brown D, Chattaway M, Dallman T, Delahay R, Kornschober C, Pietzka A, Malorny B, Petrovska L, Davies R, Robertson A, Tyne W, Weill FX, Accou-Demartin M, Williams N. The EnteroBase user’s guide, with case studies on Salmonella transmissions, Yersinia pestis phylogeny, and Escherichia core genomic diversity. Genome Research [Internet]. 2020 Jan 1 [cited 2021 Nov 13];30(1):138–52. Available from: https://genome.cshlp.org/content/30/1/138.full

23. Nguyen LT, Schmidt HA, von Haeseler A, Minh BQ. IQ-TREE: A Fast and Effective Stochastic Algorithm for Estimating Maximum-Likelihood Phylogenies. Molecular Biology and Evolution [Internet]. 2015 Jan 1 [cited 2022 Apr 30];32(1):268–74. Available from: https://academic.oup.com/mbe/article/32/1/268/2925592

24. Achtman M, Wain J, Weill FX, Nair S, Zhou Z, Sangal V, Krauland MG, Hale JL, Harbottle H, Uesbeck A, Dougan G, Harrison LH, Brisse S. Multilocus Sequence Typing as a Replacement for Serotyping in Salmonella enterica. PLOS Pathogens. 2012;8(6):e1002776.

25. Simon NC, Aktories K, Barbieri JT. Novel bacterial ADP-ribosylating toxins: structure and function. Nat Rev Microbiol [Internet]. 2014 [cited 2021 Nov 20];12(9):599. Available from: /pmc/articles/PMC5846498/

26. Afolayan AO, Oaikhena AO, Aboderin AO, Olabisi OF, Amupitan AA, Abiri O v, Ogunleye VO, Odih EE, Adeyemo AT, Adeyemo AT, Obadare TO, Abrudan M, Argimón S, David S, Kekre M, Underwood A, Egwuenu A, Ihekweazu C, Aanensen DM, Okeke IN, Abudahab K, Harste H, Muddyman D, Taylor B, Wheeler N, Donado-Godoy P, Fabian Bernal J, Arevalo A, Fernanda Valencia M, Osma Castro ECD, Ravikumar KL, Nagaraj G, Shamanna V, Govindan V, Prabhu A, Sravani D, Shincy MR, Rose S, Ravishankar KN, Ajiboye JJ, Carlos C, Lagrada ML, Macaranas PK v, Olorosa AM, Gayeta JM, Herrera EM, Molloy A, Stelling J, Vegvari C. Clones and Clusters of Antimicrobial-Resistant Klebsiella From Southwestern Nigeria. Clinical Infectious Diseases. 2021;73(Supplement_4):S308–15.

27. Petti CA, Polage CR, Quinn TC, Ronald AR, Sande MA. Laboratory medicine in Africa: A barrier to effective health care. Clinical Infectious Diseases. 2006 Feb 1;42(3):377–82.

28. Obaro SK, Hassan-Hanga F, Olateju EK, Umoru D, Lawson L, Olanipekun G, Ibrahim S, Munir H, Ihesiolor G, Maduekwe A, Ohiaeri C, Adetola A, Shetima D, Jibir BW, Nakaura H, Kocmich N, Ajose T, Idiong D, Masokano K, Ifabiyi A, Ihebuzor N, Chen B, Meza J, Akindele A, Rezac-Elgohary A, Olaosebikan R, Suwaid S, Gambo M, Alter R, Davies HD, Fey PD. Salmonella Bacteremia Among Children in Central and Northwest Nigeria, 2008–2015. Clinical Infectious Diseases. 2015 Nov 1;61(suppl_4):S325–31.

29. Popoola O, Kehinde A, Ogunleye V, Adewusi OJ, Toy T, Mogeni OD, Aroyewun EO, Agbi S, Adekanmbi O, Adepoju A, Muyibi S, Adebiyi I, Elaturoti OO, Nwimo C, Adeoti H, Omotosho T, Akinlabi OC, Adegoke PA, Adeyanju OA, Panzner U, Baker S, Park SE, Marks F, Okeke IN. Bacteremia Among Febrile Patients Attending Selected Healthcare Facilities in Ibadan, Nigeria. Clinical Infectious Diseases. 2019 Oct 30;69(Supplement_6):S466–73.

30. Akinyemi KO, Ajoseh SO, Fakorede CO. A systemic review of literatures on human Salmonella enterica serovars in Nigeria (1999-2018). The Journal of Infection in Developing Countries [Internet]. 2021 Sep 30 [cited 2022 May 19];15(09):1222–35. Available from: https://jidc.org/index.php/journal/article/view/12186

31. Ao TT, Feasey NA, Gordon MA, Keddy KH, Angulo FJ, Crump JA. Global burden of invasive nontyphoidal Salmonella disease, 2010(1). Emerg Infect Dis. 2015 Jun 1;21(6):941–9.

32. Park SE, Pham DT, Boinett C, Wong VK, Pak GD, Panzner U, Espinoza LMC, von Kalckreuth V, Im J, Schütt-Gerowitt H, Crump JA, Breiman RF, Adu-Sarkodie Y, Owusu-Dabo E, Rakotozandrindrainy R, Soura AB, Aseffa A, Gasmelseed N, Keddy KH, May J, Sow AG, Aaby P, Biggs HM, Hertz JT, Montgomery JM, Cosmas L, Olack B, Fields B, Sarpong N, Razafindrabe TJL, Raminosoa TM, Kabore LP, Sampo E, Teferi M, Yeshitela B, el Tayeb MA, Sooka A, Meyer CG, Krumkamp R, Dekker DM, Jaeger A, Poppert S, Tall A, Niang A, Bjerregaard-Andersen M, Løfberg SV, Seo HJ, Jeon HJ, Deerin JF, Park J, Konings F, Ali M, Clemens JD, Hughes P, Sendagala JN, Vudriko T, Downing R, Ikumapayi UN, Mackenzie GA, Obaro S, Argimon S, Aanensen DM, Page A, Keane JA, Duchene S, Dyson Z, Holt KE, Dougan G, Marks F, Baker S. The phylogeography and incidence of multi-drug resistant typhoid fever in sub-Saharan Africa. Nature Communications [Internet]. 2018 Dec 1 [cited 2021 Nov 24];9(1). Available from: /pmc/articles/PMC6269545/

33. Ingle DJ, Nair S, Hartman H, Ashton PM, Dyson ZA, Day M, Freedman J, Chattaway MA, Holt KE, Dallman TJ. Informal genomic surveillance of regional distribution of Salmonella Typhi genotypes and antimicrobial resistance via returning travellers. PLOS Neglected Tropical Diseases [Internet]. 2019 [cited 2021 Nov 25];13(9):e0007620. Available from: https://journals.plos.org/plosntds/article?id=10.1371/journal.pntd.0007620

34. Kingsley RA, Msefula CL, Thomson NR, Kariuki S, Holt KE, Gordon MA, Harris D, Clarke L, Whitehead S, Sangal V, Marsh K, Achtman M, Molyneux ME, Cormican M, Parkhill J, MacLennan CA, Heyderman RS, Dougan G. Epidemic multiple drug resistant Salmonella Typhimurium causing invasive disease in sub-Saharan Africa have a distinct genotype. Genome Research. 2009 Dec 1;19(12):2279–87.

35. Okoro CK, Kingsley RA, Connor TR, Harris SR, Parry CM, Al-Mashhadani MN, Kariuki S, Msefula CL, Gordon MA, De Pinna E, Wain J, Heyderman RS, Obaro S, Alonso PL, Mandomando I, MacLennan CA, Tapia MD, Levine MM, Tennant SM, Parkhill J, Dougan G. Intracontinental spread of human invasive Salmonella Typhimurium pathovariants in sub-Saharan Africa. Nature Genetics 2012 44:11. 2012 Sep 30;44(11):1215–21.

36. Kanteh A, Sesay AK, Alikhan NF, Ikumapayi UN, Salaudeen R, Manneh J, Olatunji Y, Page AJ, Mackenzie G. Invasive atypical non-typhoidal Salmonella serovars in The Gambia. Microbial Genomics [Internet]. 2021 Nov 23 [cited 2021 Nov 24];7(11):000677. Available from: https://www.microbiologyresearch.org/content/journal/mgen/10.1099/mgen.0.000677

37. Akullian A, Montgomery JM, John-Stewart G, Miller SI, Hayden HS, Radey MC, Hager KR, Verani JR, Ochieng JB, Juma J, Katieno J, Fields B, Bigogo G, Audi A, Walson J. Multi-drug resistant non-typhoidal Salmonella associated with invasive disease in western Kenya. PLOS Neglected Tropical Diseases [Internet]. 2018 Jan 1 [cited 2022 May 19];12(1):e0006156. Available from: https://journals.plos.org/plosntds/article?id=10.1371/journal.pntd.0006156

38. Park SE, Pham DT, Pak GD, Panzner U, Espinoza LMC, Kalckreuth V von, Im J, Mogeni OD, Schütt-Gerowitt H, Crump JA, Breiman RF, Adu-Sarkodie Y, Owusu-Dabo E, Rakotozandrindrainy R, Soura AB, Aseffa A, Gasmelseed N, Sooka A, Keddy KH, May J, Aaby P, Biggs HM, Hertz JT, Montgomery JM, Cosmas L, Olack B, Fields B, Sarpong N, Razafindrabe TJL, Raminosoa TM, Kabore LP, Sampo E, Teferi M, Yeshitela B, Tayeb MA el, Krumkamp R, Dekker DM, Jaeger A, Tall A, Gassama A, Niang A, Bjerregaard-Andersen M, Løfberg SV, Deerin JF, Park JK, Konings F, Carey ME, Puyvelde S van, Ali M, Clemens J, Dougan G, Baker S, Marks F. The genomic epidemiology of multi-drug resistant invasive non-typhoidal Salmonella in selected sub-Saharan African countries. BMJ Global Health [Internet]. 2021 Aug 1 [cited 2021 Nov 1];6(8):e005659. Available from: https://gh.bmj.com/content/6/8/e005659

39. Gelaw AK, Nthaba P, Matle I. Detection of Salmonella from animal sources in South Africa between 2007 and 2014. J S Afr Vet Assoc. 2018 Nov 7;89.

40. Kidanemariam A, Engelbrecht M, Picard J. Retrospective study on the incidence of Salmonella isolations in animals in South Africa, 1996 to 2006. J S Afr Vet Assoc [Internet]. 2010 [cited 2022 May 3];81(1):37–44. Available from: https://journals.co.za/doi/epdf/10.10520/EJC99859

41. Feasey NA, Dougan G, Kingsley RA, Heyderman RS, Gordon MA. Invasive non-typhoidal salmonella disease: an emerging and neglected tropical disease in Africa. The Lancet. 2012 Jun 30;379(9835):2489–99.

42. Pornsukarom S, Patchanee P, Erdman M, Cray PF, Wittum T, Lee J, Gebreyes WA. Comparative Phenotypic and Genotypic Analyses of Salmonella Rissen that Originated from Food Animals in Thailand and United States. Zoonoses and Public Health [Internet]. 2015 Mar 1 [cited 2022 May 3];62(2):151–8. Available from: https://onlinelibrary.wiley.com/doi/full/10.1111/zph.12144

43. Xu X, Biswas S, Gu G, Elbediwi M, Li Y, Yue M. Characterization of Multidrug Resistance Patterns of Emerging Salmonella enterica Serovar Rissen along the Food Chain in China. Antibiotics 2020, Vol 9, Page 660 [Internet]. 2020 Sep 30 [cited 2022 May 3];9(10):660. Available from: https://www.mdpi.com/2079-6382/9/10/660/htm

44. Hoelzer K, Switt AIM, Wiedmann M. Animal contact as a source of human non-typhoidal salmonellosis. Veterinary Research [Internet]. 2011 Feb 14 [cited 2022 May 3];42(1):1–28. Available from: https://veterinaryresearch.biomedcentral.com/articles/10.1186/1297-9716-42-34

45. Post AS, Diallo SN, Guiraud I, Lompo P, Tahita MC, Maltha J, van Puyvelde S, Mattheus W, Ley B, Thriemer K, Rouamba E, Derra K, Deborggraeve S, Tinto H, Jacobs J. Supporting evidence for a human reservoir of invasive non-Typhoidal Salmonella from household samples in Burkina Faso. PLOS Neglected Tropical Diseases [Internet]. 2019 [cited 2022 May 3];13(10):e0007782. Available from: https://journals.plos.org/plosntds/article?id=10.1371/journal.pntd.0007782

46. Aoki Y, Watanabe Y, Kitazawa K, Ando N, Hirai S, Yokoyama E. Emergence of Salmonella enterica subsp. enterica serovar Chester in a rural area of Japan. Public Health J Vet Med Sci [Internet]. 2020 [cited 2022 May 3];82(5):580–4. Available from: https://www.ncbi.nlm.nih.gov/pmc/journals/2350/

47. Reddy EA, Shaw A v., Crump JA. Community-acquired bloodstream infections in Africa: a systematic review and meta-analysis. The Lancet Infectious Diseases. 2010 Jun 1;10(6):417–32.

48. Harrois D, Breurec S, Seck A, Delauné A, le Hello S, Pardos de la Gándara M, Sontag L, Perrier-Gros-Claude JD, Sire JM, Garin B, Weill FX. Prevalence and characterization of extended-spectrum β-lactamase-producing clinical Salmonella enterica isolates in Dakar, Senegal, from 1999 to 2009. Clinical Microbiology and Infection. 2014 Feb 1;20(2):O109–16.

49. Qiu YF, Nambiar RB, Xu X bin, Weng ST, Pan H, Zheng KC, Yue M. Global Genomic Characterization of Salmonella enterica Serovar Telelkebir. Frontiers in Microbiology. 2021 Jul 29;12:2007.

50. Sokaribo AS, Perera SR, Sereggela Z, Krochak R, Balezantis LR, Xing X, Lam S, Deck W, Attah[poku S, Abbott DW, Tamuly S, White AP. A GMMA-CPS-Based Vaccine for Non-Typhoidal Salmonella. Vaccines 2021, Vol 9, Page 165 [Internet]. 2021 Feb 17 [cited 2022 Apr 29];9(2):165. Available from: https://www.mdpi.com/2076-393X/9/2/165/htm

51. Tamamura Y, Tanaka K, Uchida I. Characterization of pertussis-like toxin from Salmonella spp. that catalyzes ADP-ribosylation of G proteins. Scientific Reports 2017 7:1. 2017 Jun 1;7(1):1–13.

52. den Bakker HC, Moreno Switt AI, Govoni G, Cummings CA, Ranieri ML, Degoricija L, Hoelzer K, Rodriguez-Rivera LD, Brown S, Bolchacova E, Furtado MR, Wiedmann M. Genome sequencing reveals diversification of virulence factor content and possible host adaptation in distinct subpopulations of Salmonella enterica. BMC Genomics. 2011 Dec 22;12(1):1–11.

53. Mezal EH, Bae D, Khan AA. Detection and functionality of the CdtB, PltA, and PltB from Salmonella enterica serovar Javiana. Pathogens and Disease. 2014 Nov 1;72(2):95–103.

54. Rodriguez-Rivera LD, Bowen BM, Den Bakker HC, Duhamel GE, Wiedmann M. Characterization of the cytolethal distending toxin (typhoid toxin) in non-typhoidal Salmonella serovars. Gut Pathogens. 2015 Jul 24;7(1):1–7.

55. Chang SJ, Jin SC, Jiao X, Galan JE. Unique features in the intracellular transport of typhoid toxin revealed by a genome-wide screen. PLOS Pathogens. 2019;15(4):e1007704.

56. Marcus SL, Brumell JH, Pfeifer CG, Finlay BB. Salmonella pathogenicity islands: big virulence in small packages. Microbes and Infection. 2000 Feb 1;2(2):145–56.

57. Hensel M. Evolution of pathogenicity islands of Salmonella enterica. International Journal of Medical Microbiology. 2004 Sep 24;294(2–3):95–102.

58. Seth-Smith HMB. SPI-7: Salmonella’s Vi-Encoding Pathogenicity Island. The Journal of Infection in Developing Countries [Internet]. 2008 Aug 1 [cited 2022 May 4];2(04):267–71. Available from: https://jidc.org/index.php/journal/article/view/220

59. Shah DH, Lee MJ, Park JH, Lee JH, Eo SK, Kwon JT, Chae JS. Identification of Salmonella gallinarum virulence genes in a chicken infection model using PCR-based signature-tagged mutagenesis. Microbiology (N Y) [Internet]. 2005 Dec 1 [cited 2022 May 4];151(12):3957–68. Available from: https://www.microbiologyresearch.org/content/journal/micro/10.1099/mic.0.28126-0

60. Webber B, Borges KA, Furian TQ, Rizzo NN, Tondo EC, dos Santos LR, Rodrigues LB, do Nascimento VP. Detection of virulence genes in Salmonella Heidelberg isolated from chicken carcasses. Revista do Instituto de Medicina Tropical de São Paulo [Internet]. 2019 Jul 22 [cited 2022 May 4];61. Available from: http://www.scielo.br/j/rimtsp/a/6QcGt7kdBnqKGxbyBxXTjtn/?lang=en&format=html

61. Panzenhagen PHN, Cabral CC, Suffys PN, Franco RM, Rodrigues DP, Conte-Junior CA. Comparative genome analysis and characterization of the Salmonella Typhimurium strain CCRJ_26 isolated from swine carcasses using whole-genome sequencing approach. Letters in Applied Microbiology [Internet]. 2018 Apr 1 [cited 2022 May 4];66(4):352–9. Available from: https://onlinelibrary.wiley.com/doi/full/10.1111/lam.12859

62. Cherchame E, Guillier L, Lailler R, Vignaud ML, Jourdan-Da Silva N, le Hello S, Weill FX, Cadel-Six S. Salmonella enterica subsp. enterica Welikade: guideline for phylogenetic analysis of serovars rarely involved in foodborne outbreaks. BMC Genomics [Internet]. 2022 Dec 1 [cited 2022 May 4];23(1):1–13. Available from: https://link.springer.com/articles/10.1186/s12864-022-08439-2

63. Kingsley RA, Humphries AD, Weening EH, de Zoete MR, Winter S, Papaconstantinopoulou A, Dougan G, Bäumler AJ. Molecular and phenotypic analysis of the CS54 island of Salmonella enterica serotype Typhimurium: Identification of intestinal colonization and persistence determinants. Infection and Immunity [Internet]. 2003 Feb 1 [cited 2022 May 4];71(2):629–40. Available from: https://journals.asm.org/doi/full/10.1128/IAI.71.2.629-640.2003

64. Kombade S, Kaur N. Pathogenicity Island in <em>Salmonella</em>. Salmonella spp - A Global Challenge [Internet]. 2021 Mar 11 [cited 2022 May 4]; Available from: https://www.intechopen.com/chapters/75674

65. WHO. WHO recommends use of first typhoid conjugate vaccine [Internet]. 2018 [cited 2021 Nov 26]. Available from: https://www.who.int/news/item/03-04-2018-who-recommends-use-of-first-typhoid-conjugate-vaccine

